# Basic prediction methodology for covid-19: estimation and sensitivity considerations

**DOI:** 10.1101/2020.03.27.20045575

**Authors:** Tom Britton

## Abstract

The purpose of the present paper is to present simple estimation and prediction methods for basic quantities in an emerging epidemic like the ongoing covid-10 pandemic. The simple methods have the advantage that relations between basic quantities become more transparent, thus shedding light to which quantities have biggest impact on predictions, with the additional conclusion that uncertainties in these quantities carry over to high uncertainty also in predictions.

A simple non-parametric prediction method for future cumulative case fatalities, as well as future cumulative incidence of infections (assuming a given infection fatality risk *f*), is presented. The method uses cumulative reported case fatalities up to present time as input data. It is also described how the introduction of preventive measures of a given magnitude *ρ* will affect the two incidence predictions, using basic theory of epidemic models. This methodology is then reversed, thus enabling estimation of the preventive magnitude *ρ*, and of the resulting effective reproduction number *R*_*E*_. However, the effects of preventive measures only start affecting case fatalities some 3-4 weeks later, so estimates are only available after this time has elapsed. The methodology is applicable in the early stage of an outbreak, before, say, 10% of the community have been infected.

Beside giving simple estimation and prediction tools for an ongoing epidemic, another important conclusion lies in the observation that the two quantities *f* (infection fatality risk) and *ρ* (the magnitude of preventive measures) have very big impact on predictions. Further, both of these quantities currently have very high uncertainty: current estimates of *f* lie in the range 0.2% up to 2% ([9], [7]), and the overall effect of several combined preventive measures is clearly very uncertain.

The two main findings from the paper are hence that, a) any prediction containing *f*, and/or some preventive measures, contain a large amount of uncertainty (which is usually not acknowledged well enough), and b) obtaining more accurate estimates of in particular *f*, should be highly prioritized. Seroprevalence testing of random samples in a community where the epidemic has ended are urgently needed.

## Methods

Many papers use advanced models combined with extensive simulations to analyse and predict the outcome of the ongoing covid-19 pandemic (e.g. [3], [5] [6]). Here we complement this important literature with much simpler methods. Most likely this has the effect that estimation and prediction is less accurate than more advanced well-motivated methods. On the other hand, the methods presented here are much simpler and hence more transparent, thus allowing any reader to apply the methods to quickly obtain ball-park estimates for the most important parameters/quantities. For a more thourough statistical investigation we propose that the simple analysis presented here should be complemented with more realistic advanced models.

We consider a community that is fairly weel-mixed. It is hence more suitable for a city urban area and less for an entire country with big geographic distances - there spreading is usually more spread out in time (and space).

### Early stage predicting case fatalities and number of infected

Suppose the data at hand is the cumulative deaths Λ_*D*_(*t*), starting some calendar time *t*_0_ and ending at *t*_1_ at which still only a small community fraction has been infected. As a rule of thumb we define “early stage” by meaning that no more than 10% of the community have been infected. Mathematical theory, as well as empirical evidence for many infectious diseases, suggest that Λ_*D*_(*t*) grows approximately according to an exponential rate meaning that Λ_*D*_(*t*)*≈ const* * *e*^*rt*^ for some rate factor *r*. The exponential rate factor *r* relates to the more commonly used and easy-to-estimate doubling-time *d* by the relation *r* = ln(2)*/d*.

The growth of Λ_*D*_(*t*) will continue in the same manner until either some intervention is put in place, or until a significant community fraction have been infected. Here we consider predictions up until, say, 10% have become infected. Before this population immunity is very marginal. As a consequence, unless additional preventive measures are put in place, a very natural prediction of the cumulative number of deaths some date *t* after the final observation point *t*_1_ is given by

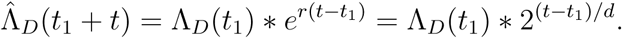

A very important quantity is how the number of infected people (n.b. not only reported!) increases over time, because this quantity carries information about how immunty builds up and when the growth rate will start declining due to partial population immunity. We hence let Λ_*I*_ (*t*) denote the total number of infected individuals up until time *t*. This cumulative number is never observed. What can be observed is the cumulative number of reported cases, but since this number is very sensitive to testing procedures which may vary over time, we refrain from using this data and stick to the more reliable reported deaths Λ_*D*_. If the *infection fatality risk f* is known, and if the typical time *s*_*d*_ between getting infected and dying (for those who die) is known (or estimated) from some other source of information, then it is straight forward to estimate Λ_*I*_(*t*) by simply back-tracking the number of “to-die” infected *s*_*d*_ days earlier and upscaling by the factor *f* :

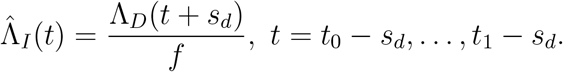

(Note that *t, t*_0_ and *t*_1_ denote calendar time whereas *s*_*d*_ is days, so if e.g. *t* = March 4 and *s*_*d*_ = 21, then *t* + *s*_*d*_ = March 25.) It is also possible to predict the cumulative number of infected later than *t*_1_ − *s*_*d*_ by using the same exponential growth as before:

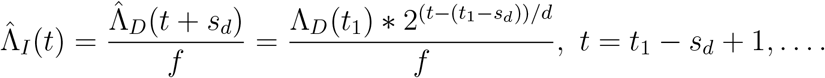

These predictions work well up until around 10% of the community have been infected, i.e. as long as 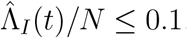, where *N* is the size of the population from which death-reports are collected.

We end this subsetion by pointing out that predictions of the number of infected are *very* sensitive to the infection fatality risk *f*. Any error in *f* carry over to prediction error multiplicatively. In many cases there is often uncertainty in *s*_*d*_, the time between infection and death and of course this duration also varies between individuals. This has the effect that time location of estimates might be off by a week or so, but the estimated numbers remain unchanged.

### Introducing preventive measures during early stage

Suppose that a new set of preventive measures are initiated on date *t*_*p*_, for example general social distancing, symptomatic people isolating, closing schools, and so on. Suppose further, that the overall effect of the preventive measures are such that the overall rate of contacts from infectious individuals to others is reduced by a factor *ρ* ∈ [0, 1].

The effect of this will be that the original *basic reproduction number R*_0_ is reduced by a factor *ρ* such that the new, *effective reproduction number R*_*E*_ is given by *R*_*E*_ = (1 − *ρ*)*R*_0_ (e.g. [4]). In order to induce how the exponential growth is affected some results from epidemic theory are needed. The exponential growth *r* is a function of the basic reproduction number *R*_0_ and the *generation time G*, where the latter is defined as the random variable describing the typical time between getting infected and infecting others. The complete relation is described by the Euler-Lotka equation (e.g. [2]) but for a wide class of epidemic models the exponential growth *r* will lie between the two relations given by

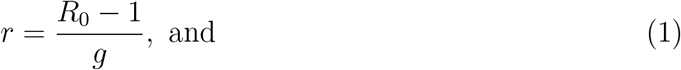

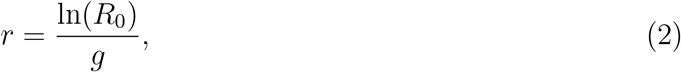

where *g* = *E*(*G*) denotes the *mean* generation time [8]. Relation (1) comes from the stochastic general epidemic model which has a highly variable generation time, whereas relation (2) comes from the Reed-Frost epidemic model for which all infections happen exactly *g* times units after getting infected, so no variation at all [2].

We now describe how to derive an expression, or rather a range of *r*_*E*_-values, to which the preventive measures will change the exponential growth to. We start by deriving an expression using relation (1). First of all, the growth rate *r* prior to preventive measures, is estimated from the cumulative reported deaths as described above. Suppose further that an estimate of *R*_0_ is available (from contact tracing, prior knowledge, or other method). Using the estimates of *r* and *R*_0_ we obtain an estimate *g*_1_ of *g* using (1). If preventive measures are introduced, then the reproduction number changes from *R*_0_ to *R*_*E*_ = (1 − *ρ*)*R*_0_. The mean generation time remains unchanged (this might not hold if enforced contact tracing is included among the introduced preventive measures). As a consequence, the new exponential growth rate satisfies

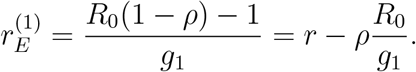

If exactly the same procedure is performed, but using relation (2) we get

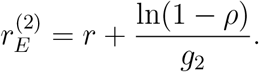

The latter expression is bigger than the former, so a natural bound for the new exponential rate *r*_*E*_, once preventive measures are put in place, is given by 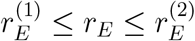. It is clear that the effect on the exponential growth rate depends to a great extent on the magnitude of the preventive effects *ρ*. It seems in general very hard to specify what overall effect *ρ* a given set of preventive measure has on reducing the rate of spread. It is hence recommended to consider a range of values rather than a fixed hypothesized value.

The procedure described above can be reversed, thus enabling estimation of the preventive effect *ρ* as follows. Suppose that the exponential growth was observed to equal *r* prior to preventive measures and that, once preventive measure have been put in place the growth rate declined to *r*_*E*_. If measured by reduced cumulative death rates we emphasize that this can only be estimated *s*_*d*_ days after the measure has been put in place (or preferably a bit later). From this it is possible to estimate *ρ* by simply inverting the procedure. As before we assume that an estimate of *R*_0_ is also available and as before we obtain two bounds using relation (1) and (2). The resulting bounds are

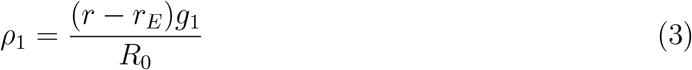

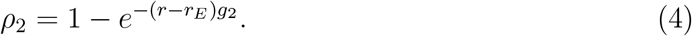

Here too, the latter expression is bigger, so a natural bound for the overall effect *ρ* is given by *ρ*_1_ ≤ *ρ* ≤ *ρ*_2_. Here we have implicitly assumed that *r*_*E*_ ≥ 0; if *r*_*E*_ < 0 then the epidemic has already started to decline. In principle the equations may still be applied (then *ρ*_2_ < *ρ*_1_) but precision quickly drops when number of newly infected is on decline.

### Prediction during the main phase of the epidemic

Predicting the epidemic during the main phase when most infections happen, so after the early stage has ended, is less straighforward and typically require a specific underlying epidemic model. To describe how this is done does not fit into the current paper relying mainly on non-parametric methods and robust basic results from epidemic theory.

For this reason we refrain from predicting the progress of the epidemic during this main phase but only give the rough picture. Once the early stage of the epidemic has passed the exponential growth rate *r* (or *r*_*E*_ in case preventive measures have been introduced) starts to decline. This goes on up until the time *t* at which the cumulative fraction infected Λ_*I*_(*t*) exceeds 1 − 1*/R*_0_ (or *R*_*E*_ in case of preventive measures). This is the time when the expected number of infections no longer exceeds one due to reduced community susceptibility. After this time the number of new infections starts dropping with time thus eventuelly leading the epidemic to a halt. The effect of introducing preventive measures reduces the initial growth rate as described in earlier sections. It also delays the timing of the peak, and particular the height of the peak, closely related the maximal healthcare burdon. The amount of peak-reduction is model dependent, but for a simple model, reducing *R*_0_ = 3 to *R*_*E*_ = 2 reduces the height of the peak by around 50%.

### Predicting final epidemic size and total number of case fatalities

We end the methodology section be given predictions of the total fraction getting infected, and perhaps even more important, the expected number of case fatalities. We assume that the same set of preventive measures (if any) remain constant through main part of the epidemic. The results are not valid if the they were put in place after a few percent had been infected, nor if they were varied during the main part of the outbreak (the prediction problem is then much more complicated and better suited for simulation studies). Suppose hence that reproduction number equals *R* (equal to *R*_0_ if no preventive measures are put in place, or otherwise *R*_*E*_). The final fraction *τ* getting infected is known to solve the equation 1 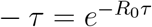 [4], a solution which has to be obtained numerically. In case the community is heterogeneous in one or several ways, the final fraction getting infected is usually 5-20% smaller than this solution. So, given that the reproduction number is known the final fraction getting infected is well-predicted. The estimate is of course sensitive to *R*, but if *R ≥* 2 then *τ ≥* 75%, so it is not too sensitive to exakt values of *R* in case it is above 2 or so.

If the infection fatality risk *f* is known, then the predicted number of case fatalities is *N* * *f* \**τ*, where as before *N* denotes the population size. This estimate is very sensitive to *f*. Since *f* is typically very small, possible values may well vary by a factor 10, implying that predictions of case fatalities will also vary by a factor 10.

## Illustrations

We now illustrate our simple methodology to realistic covid-19 settings. We start be making predictions assuming no preventive measures whatsoever. In most European countries the doubling time prior to prevntive measures lie very close to *d* = 3 days. A common estimate of *R*_0_ equals 2.5, but estimates seem to go up so we use *R*_0_ = 3 in our illustration. As our last cumulative death number we pick *t*_1_ = March 24 and assume that the urban area of interest has a population of *N* = 3 million and by March 24 had experienced 50 case fatalities (Λ_*D*_(*t*_1_) = 50). A prediction for the cumulative number of deaths by *t* = April 3 (10 days later) hence equals 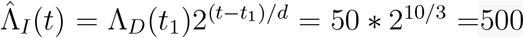. We assume that *s*_*d*_, the typical time from getting infected to dying (assuming the individual dies from covid-19), equals 21 days, and that the infection fatality risk equals *f* = 0.3%. With these assumptions, the estimate of how many people that had been infected by *t* = March 3 (3 weeks earlier) equals 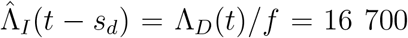. If we want to estimate how infected there are at *t* = March 15, this number has to be scaled up using the doubling times: Λ_*I*_(*t*) = Λ_*D*_(*t*_1_) * 2^(15−3)*/*3^*/f* = 267 000, thus approaching the limit of 10% where predictions should stop using the current method. Nedless to say, there is of course quite a lot of uncertainty in such a prediction: first we estimated the number of infected 3 weeks back using case fatalities of present time and then projected this number three weeks forward in time to present using the doubling times.

If *R*_0_ = 3 and the doubling time equals *d* = 3 corresponding to an exponential growth rate of *r* = ln(2)*/*3 = 0.23. If the magnitude of the overall effect of preventive measures equals *ρ* = 1*/*3, then using relation (1) gives a mean generation time *g*_1_ = (*R*_0_ − 1)*/r* = 8.7 days. The new exponential growth rate after preventive measure are put in place hence equals 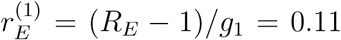. If instead relation (2) is used, the mean generation time is *g*_2_ = ln(*R*_0_)*/r* = 4.5 days. As a consequence, the new exponential growth rate assuming relation (2) equals 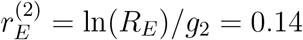. From the two bounds we hence conclude that *r*_*E*_ should satisfy 0.11 *≤ r*_*E*_ *≤* 0.14.

We now include preventive measures. We still assume *R*_0_ = 3 and a doubling time of *d* = 3 days without preventions. Then preventive measures are put in place, resulting in a prolonged doubling time of case fatalities starting (around) *s*_*d*_ = 21 days later. Suppose that the new observed doubling time equals *d*_*E*_ = 5 days. These two doubling times correspond to the exponential rates *r* = 0.23 and *r*_*E*_ = ln(2)*/*5 = 0.14. If we first assume relation (1) to hold, this reduced exponential rate gives a lower bound the magnitude *ρ* given by *ρ*_1_ = (*r* − *r*_*E*_)*g*_1_*/R*_0_ = 0.27. If we instead consider relation (2) we obtain the upper bound of the magnitude as 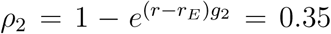. It hence follows that the magnitude *ρ* of the preventive measure giving rise to an increased doubling time from *d* = 3 to *d*_*E*_ = 5 days is estimated to lie somewhere in the interval (0.27, 0.35). The effective reproduction *R*_*E*_ is hence estimated to lie between *R*_0_(1 − 0.35) = 1.93 and *R*_0_(1 − 0.27) = 2.19, so a substantial reduction.

We end by predicting the final fraction infected, and the total number of case fatalities, at the end of the outbreak. This is possible from simply knowing the reproduction number, as described in the method section. For instance, if *R*_0_ = 3 the final size equation gives *τ* = 94%. If preventive measures with magnitude *ρ* = 1*/*3 are put in place during the early stage and kept all trough the outbreak, then the predicted final size instead equals *τ*_*E*_ = 78% In the more likely case of a heterogeneous community the fractions should be deflated by about 20%. Similarly, the predicted total number of case fatalities is *Nfτ* = and *Nfτ*_*E*_ respectively. Since communities typically are heterogeneous better estimates would be to deflate these numbers by about 20%.

## Conclusions

One of our main conclusions is that any prediction of the number of infected, using data from cumulative case fatalities, rely heavily on accurate knowledge of the infection fatality risk *f*. Without accurate knowledge of *f*, the prediction of the total case fatality once the epidemic outbreak is over, is equipped with a similar multiplicative uncertainty. It is hence of utmost importance to obtain more accurate estimates of *f*. The best way to obtain such estimates, now that seroprevalence tests are becoming available, is to conduct tests on a random sample of a community in which the outbreak is (close to) over. *f* is then simply estimated as the ratio of the total number of case fatalities and the estimated population number of people testing positive.

Further, combinations of preventive measures typically have a very uncertain overall magnitude *ρ*, and it was shown how this uncertainty carries over to high uncertainty in estimates of the effective reproduction number *R*_*E*_ and the corresponding decline in growth rate *r*_*E*_. However, it was also illustrated how to estimate *ρ* if cumulative deaths are collected a bit longer than *s*_*d*_ days ahead before estimating.

As mentioned earlier, the main idea with the present paper is to keep methods simple enough to make the methodology transparent. We highly recommend to complement this simple analysis with more detailed models which might give better precision (at the price of not being transparent).

The methodology presented used cumulative reported deaths as the main data source. A problem with this data is that there is a delay of about 3 weeks from getting infected to dying (for those who die). As a consequence, there is a more than 3 week delay before effects of interventions may be estimated. The same methodology can be used for data on cumulative number of patients having received intensive care (IC) treatment. The time from getting infected to the start of IC treatment (for those who need it) is around 2 weeks, thus making the delay shorter. This must be weighted against the often lower quality data for IC treatment as compared to reported deaths.

Finally we stress that in the current paper we have not at all treated statistical uncertainty of the estimators and predictions. Deriving standard errors for these quantities is far from easy, and are often also model-dependent. Such methodology is hence beyong the scope of the present paper where we focus on simple methodology. We also believe the uncertainty in *f* and *ρ* is orders of magnitude larger than statistical uncertainty.

## Data Availability

No data is used

